# Hematopoietic and Lung Platelet Biogenesis as a Prognostic Indicator in Idiopathic Pulmonary Fibrosis (IPF)

**DOI:** 10.1101/2022.04.04.22273424

**Authors:** Shigeki Saito, Cheng Han H. Chung, Alex Jacob, Nebil Nuradin, Amy E. Meyer, Toshie Saito, Haoran Yang, Jay K. Kolls, Victor J. Thannickal, Yao-Zhong Liu, Joseph A. Lasky

## Abstract

**Rationale and objectives:** The role of human lung megakaryocytes in homeostasis and their dynamics in disease states remain unknown. We sought to investigate whether megakaryocyte/platelet gene signatures are altered in IPF.

**Methods:** We analyzed publicly available transcriptome datasets of lung tissue, bronchoalveolar lavage (BAL) cells, and peripheral whole blood from IPF patients and healthy controls. Enrichment of megakaryocyte and platelet gene signatures in those datasets were estimated using xCell, a novel computational method. Furthermore, we analyzed whether mean platelet volume (MPV) and platelet counts in peripheral blood are associated with lung transplant-free survival in our IPF cohort.

**Results:** In lung tissue, megakaryocyte gene signature enrichment scores were significantly lower in IPF than in controls. In BAL cells, platelet gene signature enrichment scores were significantly lower in IPF than in controls, and lower platelet scores were associated with lower lung transplant-free survival in IPF. In contrast, in blood, megakaryocyte scores were significantly higher in IPF than in controls, and higher megakaryocyte scores were associated with lower disease progression-free survival in IPF. Furthermore, higher MPV was associated with lower transplant-free survival in our IPF cohort, independent of age, sex, forced vital capacity (FVC), and diffusing capacity of the lung for carbon monoxide (DLCO).

**Conclusions:** In IPF, megakaryocyte/platelet gene signatures were altered in a compartment-specific manner. Moreover, those signatures and MPV in blood were associated with important clinical outcomes such as transplant-free survival. These findings provide new insights into altered megakaryocyte/platelet biogenesis in IPF and suggest the potential utility of megakaryocyte/platelet-based biomarkers in IPF.

## Introduction

A recent study indicated that, in mice, the lung harbors megakaryocytes and is a site of platelet biogenesis (1). The presence of megakaryocytes has also been reported in human lungs (2, 3); however, the characteristics and role of lung megakaryocytes in homeostatic conditions, as well as chronic lung diseases such as idiopathic pulmonary fibrosis (IPF), are not well understood.

We investigated whether megakaryocyte/platelet gene signatures are altered in IPF by analyzing publicly available transcriptome datasets with xCell (4), a novel computational method. We further sought to determine whether platelet counts and mean platelet volume (MPV) in peripheral blood, two platelet parameters routinely measured in clinical practice (as a part of complete blood count [CBC]), are associated with lung transplant-free survival in our own IPF cohort.

## Methods

We analyzed the following NCBI Gene Expression Omnibus (GEO) transcriptome datasets from IPF patients and healthy controls: lung tissue (GSE47460_GPL6480, GSE47460_GPL14550), bronchoalveolar lavage (BAL) cells (GSE70867), and peripheral whole blood (GSE33566, GSE93606). We generated gene enrichment scores for megakaryocytes and platelets with xCell, a computational method that assesses enrichment of individual cell types based on gene expression profile, and compared the scores between IPF patients and controls using unpaired t-test or Mann-Whitney test. In GSE70867 and GSE93606 (where survival data were available), we compared event-free survival between patients with higher enrichment scores (above the median) and patients with lower enrichment scores (below the median) using a log-rank test.

We also analyzed data from our own Tulane IPF cohort to determine whether platelet counts and MPV in peripheral blood are associated with lung transplant-free survival. We performed Cox proportional hazards regression analysis, adjusting for age, sex, forced vital capacity (FVC), and diffusing capacity of the lung for carbon monoxide (DLCO). This study was approved by the Tulane University Institutional Review Board. A p-value <0.05 was considered statistically significant.

## Results

In lung tissue, samples from IPF patients had significantly lower megakaryocyte enrichment scores than controls (i.e., IPF lung transcriptome was significantly less enriched with a megakaryocyte gene signature than the control lung transcriptome) (Figure 1A). Platelet scores were not significantly different between IPF lung transcriptome and control lung transcriptome. In BAL cells, IPF patients had significantly lower platelet scores than controls, and IPF patients with lower platelet scores had significantly worse lung transplant-free survival rates than IPF patients with higher scores (log-rank test p<0.0001, hazard ratio=2.190 [95% CI: 1.476–3.249]) (Figure 1B). In contrast, in peripheral whole blood, IPF patients had significantly higher megakaryocyte scores than controls, and IPF patients with higher megakaryocyte scores had significantly worse disease progression-free survival (i.e., survival without significant [>10%] decline in FVC over six months) than IPF patients with lower megakaryocyte scores (log-rank test p=0.0096, hazard ratio=2.535 [95% CI: 1.254–5.128]) (Figure 1C). Platelet scores were not significantly different between IPF blood transcriptome and control blood transcriptome.

**Figure 1.**
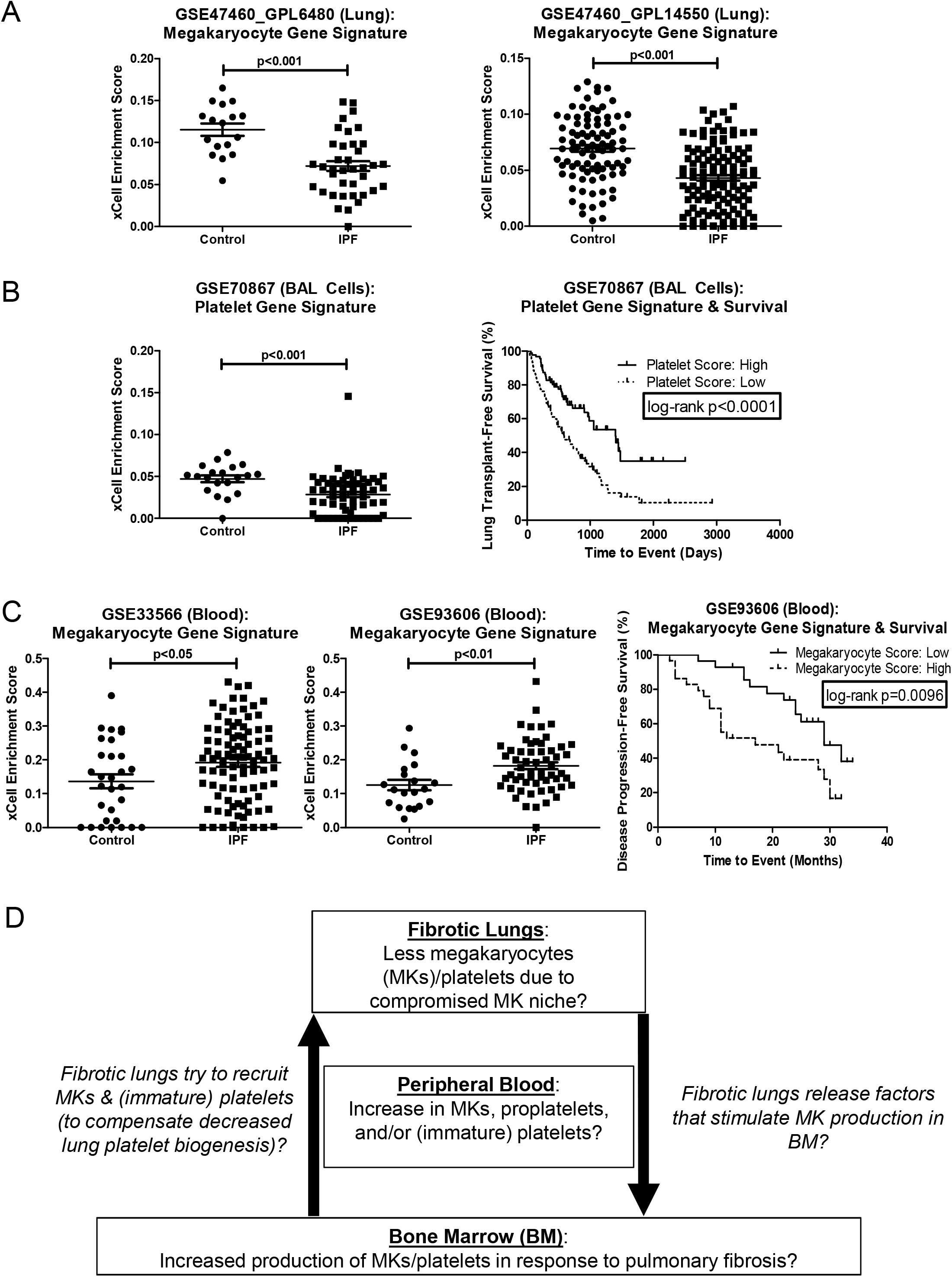
Megakaryocyte/platelet gene signatures were altered in IPF in a compartment-specific manner. We analyzed IPF transcriptome datasets available in NCBI Gene Expression Omnibus (GEO). We generated gene enrichment scores for megakaryocytes and platelets with xCell (https://xcell.ucsf.edu/), a novel computational method that assesses enrichment of individual cell types based on gene expression profile. (A) In lung transcriptome (GSE47460_GPL6480, GSE47460_GPL14550), IPF patients had significantly lower megakaryocyte enrichment scores than controls (who underwent surgery for the investigation of a nodule but had no chronic lung disease by CT or pathology). (B) In BAL cell transcriptome (GSE70867), IPF patients had significantly lower platelet enrichment scores than controls (*Left*: Freiburg cohort only, for which controls were available). IPF patients with lower platelet scores (i.e., lower than median) had significantly worse lung transplant-free survival than IPF patients with higher platelet scores (i.e., higher than median) (log-rank test p<0.0001, hazard ratio=2.190 [95% CI: 1.476–3.249]) (*Right*: overall cohort [=Freiburg, Siena, and Leuven cohort]). (C) In peripheral whole blood transcriptome (GSE33566, GSE93606), IPF had significantly higher megakaryocyte scores than age- and sex-matched healthy controls (*Left, middle*). IPF patients with higher megakaryocyte scores (above the median) had worse disease progression-free survival* than IPF patients with lower megakaryocyte scores (below the median) (log-rank test p=0.0096, hazard ratio=2.535 [95% CI: 1.254–5.128]) (*Right*). *Disease progression-free survival was defined as survival without significant [>10%] decline in FVC over six months. (D) Working hypothesis (proposed schema): Altered megakaryocyte/platelet biogenesis in IPF

In our Tulane IPF cohort (n=184; Table 1), univariate analysis revealed that higher MPV (but not higher platelet count) was significantly associated with worse lung transplant-free survival (p=0.042). More importantly, multivariate analysis revealed that higher MPV (but not higher platelet count) was significantly associated with worse lung transplant-free survival, independent of age, sex, FVC, and DLCO (p=0.031) (Multivariate analysis 1, 2). Furthermore, higher MPV was significantly associated with worse lung transplant-free survival, independent of age, sex, FVC, DLCO, and platelet counts (p=0.012) (Multivariate Analysis 3).

**Table 1.** Survival analysis in our Tulane IPF cohort. IPF patients followed at Tulane Medical Center or University Medical Center New Orleans were enrolled in this study (n= 184). The diagnosis of IPF was made according to the international guideline (10). Data on age, sex, and platelet counts were available from all 184 patients. Data on FVC, DLCO, MPV, and monocyte counts were available from 177, 169, 165, and 167 patients, respectively. We performed univariate analysis with variables that have been shown to be associated with survival in multiple studies, then performed multivariate analysis with variables with p<0.20 in univariate analysis. Univariate analysis revealed that higher MPV (but not higher platelet counts) was significantly associated with worse lung transplant-free survival (p=0.042, hazard ratio=1.210 [95% CI: 1.007–1.453]). Multivariate analysis revealed that higher MPV (but not higher platelet counts) was significantly associated with worse lung transplant-free survival, independent of age, sex, FVC, and DLCO (p=0.031, hazard ratio=1.235 [95% CI: 1.020–1.496]) (Multivariate analysis 1, 2). Moreover, higher MPV (but not higher platelet counts) was significantly associated with worse lung transplant-free survival, independent of age, sex, FVC, DLCO, and platelet counts (p=0.012, hazard ratio=1.291 [95% CI: 1.056–1.577]) (Multivariate analysis 3). * IQR = interquartile range; FVC = forced vital capacity; DLCO = diffusing capacity of the lung for carbon monoxide; MPV = mean platelet volume

| Clinical characteristics |  |  |
| --- | --- | --- |
| Age (years): median (IQR) | 69.6 (64.5–75.7) |  |
| Sex: Male, n (%) | 129 (70.1) |  |
| FVC (% predicted): median (IQR) | 69.0 (55.7–81.0) |  |
| DLCO (% predicted): median (IQR) | 45 (37.5–55.5) |  |
| MPV (femtoliters): median (IQR) | 9.90 (8.90–10.6) |  |
| Platelet Counts (per microliter): median (IQR) | 229 (190–264) |  |
| Monocyte Counts (×10<sup>9</sup> per liter): median (IQR) | 0.670 (0.55–0.84) |  |
| Cox proportional hazards regression model |  |  |
|  | P value | Hazard Ratio (95% Confidence Interval) |
| Univariate Analysis |  |  |
| Age | 0.006 | 1.037 (1.010–1.064) |
| Sex (male compared to female) | 0.162 | 1.446 (0.863–2.425) |
| FVC (% predicted) | <0.001 | 0.970 (0.957–0.984) |
| DLCO (% predicted) | <0.001 | 0.961 (0.944–0.979) |
| MPV | 0.042 | 1.210 (1.007–1.453) |
| Platelet Counts | 0.114 | 1.003 (0.999–1.006) |
| Monocyte Counts | 0.502 | 1.435 (0.500–4.116) |
| Multivariate Analysis 1 |  |  |
| Age | 0.467 | 1.011 (0.982–1.040) |
| Sex (male compared to female) | 0.236 | 1.467 (0.779–2.763) |
| FVC (% predicted) | 0.031 | 0.982 (0.966–0.998) |
| DLCO (% predicted) | 0.002 | 0.969 (0.950–0.989) |
| MPV | 0.031 | 1.235 (1.020–1.496) |
| Multivariate Analysis 2 |  |  |
| Age | 0.172 | 1.020 (0.992–1.048) |
| Sex (male compared to female) | 0.477 | 1.245 (0.680–2.282) |
| FVC (% predicted) | 0.007 | 0.980 (0.965–0.994) |
| DLCO (% predicted) | 0.009 | 0.976 (0.959–0.994) |
| Platelet Counts | 0.147 | 1.003 (0.999–1.007) |
| Multivariate Analysis 3 |  |  |
| Age | 0.454 | 1.011 (0.982–1.040) |
| Sex (male compared to female) | 0.122 | 1.670 (0.872–3.200) |
| FVC (% predicted) | 0.026 | 0.982 (0.967–0.998) |
| DLCO (% predicted) | 0.003 | 0.971 (0.952–0.990) |
| MPV | 0.012 | 1.291 (1.056–1.577) |
| Platelet Counts | 0.075 | 1.004 (1.000–1.009) |

## Discussion

Our analysis revealed that megakaryocyte/platelet gene signatures were altered in IPF in a compartment-specific manner. Based on our findings, one could speculate that IPF lungs harbor fewer (healthy) megakaryocytes due to their altered megakaryocyte niche and that megakaryocytes are actually decreased in IPF lungs. So far, published single-cell RNA-seq analyses have failed to identify a cluster of lung megakaryocytes in fibrotic lung tissue (5, 6). This may be because lung megakaryocytes are too rare or fragile to be isolated without any enrichment or a special protocol optimized for megakaryocytes.

There are a few possible explanations for less enrichment of megakaryocyte gene signature in IPF *lung* transcriptome and more enrichment of megakaryocyte gene signature in IPF *blood* transcriptome compared to control counterparts (Figure 1D). One possibility is that decreased megakaryocyte/platelet biogenesis in IPF lungs induces megakaryocyte/platelet biogenesis in bone marrow. For example, bone marrow may be producing more megakaryocytes and/or releasing more megakaryocytes into the circulation in response to factors released by fibrotic lungs. The second possibility is that megakaryocytes cannot remain in the altered niche within IPF lungs and therefore escape into the circulation. The third possibility is that fibrotic lungs are “educating” circulating platelets and modifying their transcriptome, as shown in previous works on “tumor-educated platelets” (7).

There are notable limitations to our study. One major limitation is that *in silico* prediction/estimation of gene signature enrichment for various types of cells in tissues is still far from perfect. Another limitation is that a megakaryocyte gene signature derived from bone marrow megakaryocytes is unlikely to be fully applicable to lung megakaryocytes, because lung megakaryocytes are transcriptionally (and phenotypically) distinct from bone marrow megakaryocytes. For example, lung megakaryocytes have higher expression of MHC class II and other molecules related to antigen presentation compared to bone marrow megakaryocytes (1, 8, 9). Furthermore, the prognostic utility of platelet gene signature in BAL cells, megakaryocyte gene signature in blood, and MPV in blood needs to be confirmed in other validation cohorts. Lastly, our study did not answer the following important questions: 1) Do megakaryocytes and/or platelets contribute to the pathogenesis of IPF? 2) Is the human lung a reservoir of megakaryocyte progenitors and a source of platelet biogenesis, as observed in mouse lungs? Further studies are needed to address these questions.

In conclusion, megakaryocyte/platelet gene signatures are altered and may have prognostic value in IPF. Moreover, MPV, a platelet parameter routinely measured in clinical practice, may be a simple yet novel prognostic biomarker in IPF. Further studies are needed to elucidate megakaryocyte/platelet biogenesis and to validate prognostic utility of megakaryocyte/platelet gene signatures as well as MPV in IPF.

## Data Availability

All data produced in the present work are contained in the manuscript.

## References

1. Lefrançais E, Ortiz-Muñoz G, Caudrillier A, Mallavia B, Liu F, Sayah DM, Thornton EE, Headley MB, David T, Coughlin SR, Krummel MF, Leavitt AD, Passegué E, Looney MR. The lung is a site of platelet biogenesis and a reservoir for haematopoietic progenitors. Nature 2017;544:105–109.

2. Rapkiewicz AV, Mai X, Carsons SE, Pittaluga S, Kleiner DE, Berger JS, Thomas S, Adler NM, Charytan DM, Gasmi B, Hochman JS, Reynolds HR. Megakaryocytes and platelet-fibrin thrombi characterize multi-organ thrombosis at autopsy in COVID-19: A case series. EClinicalMedicine 2020;24:100434.

3. Valdivia-Mazeyra MF, Salas C, Nieves-Alonso JM, Martín-Fragueiro L, Bárcena C, Muñoz-Hernández P, Villar-Zarra K, Martín-López J, Ramasco-Rueda F, Fraga J, Jiménez-Heffernan JA. Increased number of pulmonary megakaryocytes in COVID-19 patients with diffuse alveolar damage: an autopsy study with clinical correlation and review of the literature. Virchows Arch 2021;478:487–496.

4. Aran D, Hu Z, Butte AJ. xCell: digitally portraying the tissue cellular heterogeneity landscape. Genome Biol 2017;18:220.

5. Adams TS, Schupp JC, Poli S, Ayaub EA, Neumark N, Ahangari F, Chu SG, Raby BA, DeIuliis G, Januszyk M, Duan Q, Arnett HA, Siddiqui A, Washko GR, Homer R, Yan X, Rosas IO, Kaminski N. Single-cell RNA-seq reveals ectopic and aberrant lung-resident cell populations in idiopathic pulmonary fibrosis. Sci Adv 2020;6:eaba1983.

6. Habermann AC, Gutierrez AJ, Bui LT, Yahn SL, Winters NI, Calvi CL, Peter L, Chung M-I, Taylor CJ, Jetter C, Raju L, Roberson J, Ding G, Wood L, Sucre JMS, Richmond BW, Serezani AP, McDonnell WJ, Mallal SB, Bacchetta MJ, Loyd JE, Shaver CM, Ware LB, Bremner R, Walia R, Blackwell TS, Banovich NE, Kropski JA. Single-cell RNA sequencing reveals profibrotic roles of distinct epithelial and mesenchymal lineages in pulmonary fibrosis. Sci Adv 2020;6:eaba1972.

7. In ‘t Veld Sgjg, Wurdinger T. Tumor-educated platelets. Blood 2019;133:2359–2364.

8. Pariser DN, Hilt ZT, Ture SK, Blick-Nitko SK, Looney MR, Cleary SJ, Roman-Pagan E, Saunders J, Georas SN, Veazey J, Madere F, Santos LT, Arne A, Huynh NP, Livada AC, Guerrero-Martin SM, Lyons C, Metcalf-Pate KA, McGrath KE, Palis J, Morrell CN. Lung megakaryocytes are immune modulatory cells. J Clin Invest 2021;131:137377.

9. Yeung AK, Villacorta-Martin C, Hon S, Rock JR, Murphy GJ. Lung megakaryocytes display distinct transcriptional and phenotypic properties. Blood Adv 2020;4:6204–6217.

10. Raghu G, Remy-Jardin M, Myers JL, Richeldi L, Ryerson CJ, Lederer DJ, Behr J, Cottin V, Danoff SK, Morell F, Flaherty KR, Wells A, Martinez FJ, Azuma A, Bice TJ, Bouros D, Brown KK, Collard HR, Duggal A, Galvin L, Inoue Y, Jenkins RG, Johkoh T, Kazerooni EA, Kitaichi M, Knight SL, Mansour G, Nicholson AG, Pipavath SNJ, et al. Diagnosis of Idiopathic Pulmonary Fibrosis. An Official ATS/ERS/JRS/ALAT Clinical Practice Guideline. Am J Respir Crit Care Med 2018;198:e44–e68.

